# The Maldistribution of Pediatric Providers as a Potential Driver of Reduced Educational Outcomes

**DOI:** 10.1101/2021.05.24.21257733

**Authors:** Jessica C. Boyle, Benjamin W. Domingue

## Abstract

**Objective:** Despite evidence of a strong bidirectional connection between educational achievement and health, few studies have examined the link between these intertwining forces on a national level. This study takes advantage of a new population-level dataset to explicitly link child health access to academic outcomes in nearly every U.S. school district.

**Methods:** National data were used to construct and link district-level measures of child health access to district-level measures of third-grade achievement. Specifically, location data for over 256,000 practicing pediatricians and family physicians were linked to achievement data from 12,296 school districts. We include district-level rates of uninsured children as an additional measure of child health access.

**Results:** First, physician supply is unequally distributed across districts and their student populations. Second, districts that had higher physician supply tended to have higher test scores. This relationship is most pronounced for districts with relatively few pediatricians and family physicians. While the rate of uninsured children is largely correlated with community socioeconomic status, physician supply appears to operate independently of this measure.

**Conclusion:** Early childhood health and wellbeing are linked to cognitive performance and achievement in school. We provide evidence to illustrate an aspect of this relationship: children with less access to healthcare providers also do less well in school. The specific patterning of this finding suggests a need to reconsider how availability and access to pediatricians and family physicians is currently configured. Future research should examine whether a redistribution of the existing physician workforce could result in a net academic benefit for students.

## Introduction

Decades of research in child development have confirmed the importance of early childhood in shaping life outcomes, and education has been one of the processes by which scholars, practitioners, and policymakers have sought to improve these outcomes.^1,2^ Advances in our understanding of child development have led to calls for increased interdisciplinary collaboration to guide the future of early childhood policy.^3,4^ Education researchers likewise acknowledge that gaps in achievement along lines of income and race/ethnicity are attributable to structural inequalities in out-of-school factors and are evident very early in life^5–9^; prominent leaders in education have pointed out that children spend 80% of their waking hours outside of the classroom, and that it is necessary to meet students’ basic needs in order to ensure they arrive in the classroom ready to learn.^10,11^ To meet these needs, there is a growing movement to better integrate health and educational services.^12,13^

Contemporaneous advancement in our geographic understanding of both health and education inequities have resulted in parallel yet disconnected findings on the spatial organization of child wellbeing and opportunity. Recent innovations in data collection have enabled researchers to examine geographic disparities in health and educational access more comprehensively than ever before.^14,15^ The literature detailing these geographic disparities across the health and education sectors tell similar stories: when it comes to health and educational outcomes, place matters.^6,16–22^ However, despite the intertwining role of education and health in overall child wellbeing, little research has linked these disparities across space. We thus focus on the following questions: Are places that have low levels of child health access generally the same places that have low levels of quality educational access? Do places with more supply tend to have specific sociodemographic characteristics? And finally, to what extent are these disparities associated with one another?

The geographic distribution of physicians is one of the myriad ways that place influences health access. A 1997 county-level analysis found that pediatricians were less evenly distributed than physicians overall, and that this inequality grew over time; furthermore, higher pediatrician-to-child-population ratios were associated with higher state per capita income and higher numbers of residency slots.^23^ Another analysis found that while the general pediatrician and family physician workforces expanded faster than the child population between 1996 and 2006, there were enormous disparities in the 2006 per capita supply of such physicians: while 20% of the child population lived in local primary care markets with less than 710 children per physician, another 20% of the population lived in local markets of over 4,400 children per physician. Another 1 million children lived in areas with no local child physician whatsoever.^24^ While these papers are informative, they do not provide an up-to-date description of the distribution of pediatricians and family physicians, nor do they describe the populations present in their local communities.

We examine how physicians trained as either pediatricians or family physicians (who we refer to in this paper as *pediatric providers*) are distributed across U.S. school districts, investigate whether health access and quality educational access intersect across place, and explore how these highly influential systems potentially contribute to each other. We generate district-level measures of physician supply and the rate of uninsured children to represent child health access; these factors are linked to beneficial health^25–32^ or educational^33–35^ outcomes for children. Using these data, we first examine the distribution of pediatric providers across U.S. school districts. We then investigate district-level characteristics associated with supply. Finally, we analyze associations between this distribution and local levels of achievement.

## Methods

### Data

The education data source for this paper is the Stanford Education Data Archive (SEDA, version 4.0), which uses nearly 430 million standardized test scores from all U.S. public school students in grades 3 to 8 to construct measures of academic achievement for every community in America between academic years 2008-2009 and 2017-2018. SEDA assessment data is drawn from the ED*Facts* database at the U.S. Department of Education, then linked to a common scale using the National Assessment of Educational Progress (NAEP); this enables researchers to compare student achievement across grades, states, and years.^36^ From SEDA we use third-grade test scores as our achievement measure; this is to focus our attention on early childhood, the period for which we have the most evidence linking health and educational achievement. In order to interpret the magnitude of this measure, the average U.S. student’s score improves by one-third of a standard deviation (SD) per grade. Thus, a district where average test scores are 0.33 SD higher than the national average is roughly one grade level ahead of the national average for that grade.

SEDA provides estimates of each district’s average socioeconomic status (SES) using the NCES Education Demographic and Geographic Estimates (EDGE) program data, which tabulates American Community Survey (ACS) data within geographic school district boundaries. The ACS and EDGE data are reported as 5-year averages and SEDA uses the 2005-2009 through 2014-2018 waves of EDGE data. The SES measure is constructed by taking the first principal component of six variables reported in the EDGE data: median family income, proportion of adults with a bachelor’s degree or higher, household poverty rates, proportion of adults that are unemployed, proportion of households receiving SNAP benefits, and proportion of households with children that are headed by a single mother. We use an Empirical Bayes “shrunken” estimate of the SES composite in our analyses. The district-level racial/ethnic composition measure is derived from school-level covariate data that is drawn from the Common Core of Data (CCD), which provides the percentage of students eligible for free or reduced-price lunch and the racial/ethnic composition of students in each school.

We construct a measure of district pediatric provider supply by creating a physician-to-child-population ratio (PCPR) for every school district in our sample. We first generate a count of pediatric providers in every district by accessing the American Medical Association Physician Masterfile, which provides the practice location of every active physician in the United States.

We use the healthcare provider taxonomy codes to restrict physician observations to pediatricians and family physicians only, resulting in over 256,000 physicians whose addresses are then geocoded onto a geographic school district shapefile. The resulting data provides the count of pediatric providers who practice within the boundaries of every U.S. geographic school district. To convert this into a physician-to-child-population ratio, we access child population estimates from two sets of ACS 5-Year Data. This variable provides an estimate of the child population in every U.S. geographic school district through multi-year sampling, and we combine 2009-2013 data with 2014-2018 data to reduce sampling error and produce more reliable estimates. Using the district physician counts and district child population estimates, we generate a ratio representing the number of pediatric providers per 1,000 kids.

To construct the rate of uninsured children in each school district, we access ACS 5-Year district health insurance data for years 2009-2013 and 2014-2018. We restrict the sample for each file to the pediatric population, producing a count of uninsured children in every district across the country. We divide this number by the child population estimate in each respective file to produce the rate of uninsured children. Finally, we average the 2009-2013 and 2014-2018 insurance rates together to reduce sampling error and produce more reliable estimates. To account for uncertainty in our insurance measure, particularly in districts with smaller populations, we generate Empirical Bayes estimates for the district rate of uninsured children; these estimates are used as predictors. We remove 47 observations with child population measures that are over 30% noise and exclude an additional 45 observations with a PCPR over 35 and a rate of uninsured children over 44%. This results in a sample of 12,296 school districts. The supplemental information (SI) contains sensitivity analyses conducted on data that includes excluded districts; results are similar.

### Models

We fit a set of regression models to estimate the associations between district test score outcomes and district health indicators. The models take the form

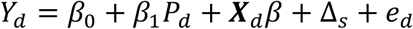

where *Y*_*d*_ is the estimated third-grade achievement in district *d*, averaged across subjects (mathematics and English Language Arts) and years; *P*_*d*_ is district physician-to-child-population ratio; and ***X***_*d*_ is a vector of district covariates (community socioeconomic status, percentage of students that are White, and the rate of uninsured children). We include a fixed effect for the state, Δ_*S*_, to eliminate any bias introduced by unobservable state-level characteristics.

## Results

### The Distribution of Pediatric Providers

There are over 256,000 pediatricians and family physicians distributed across 8,657 of the 12,296 school districts included in this sample (3,639 districts have no pediatric provider). The average district has 2.5 pediatric providers per 1,000 children. Looking strictly at pediatricians—rather than the combined total of pediatricians and family physicians—we find that there are over 80,000 pediatricians distributed across just 4,332 of 12,296 school districts— in other words, nearly 8,000 U.S. districts have no pediatrician within its boundaries. The average district has 0.43 pediatricians per 1,000 children.

We can contrast the patterning of pediatric provider density across districts with other district-level features. While the density of pediatric providers is highly associated with the district’s rurality, it is relatively unassociated with other district features. Figure 1 shows correlations between key district features. There is a correlation of –0.29 between PCPR and being a rural district; correlation between all other variables is less than 0.09 in magnitude. Note that the density of pediatric providers is unassociated with district-level SES (r=-0.007, p=0.417). In general, these unadjusted associations between physician density and other features are quite low, especially in comparison to the generally larger associations observed between district SES and racial/ethnic composition. In contrast, the rate of uninsured children is moderately correlated with SES and percentage of White students.

**Figure 1.**
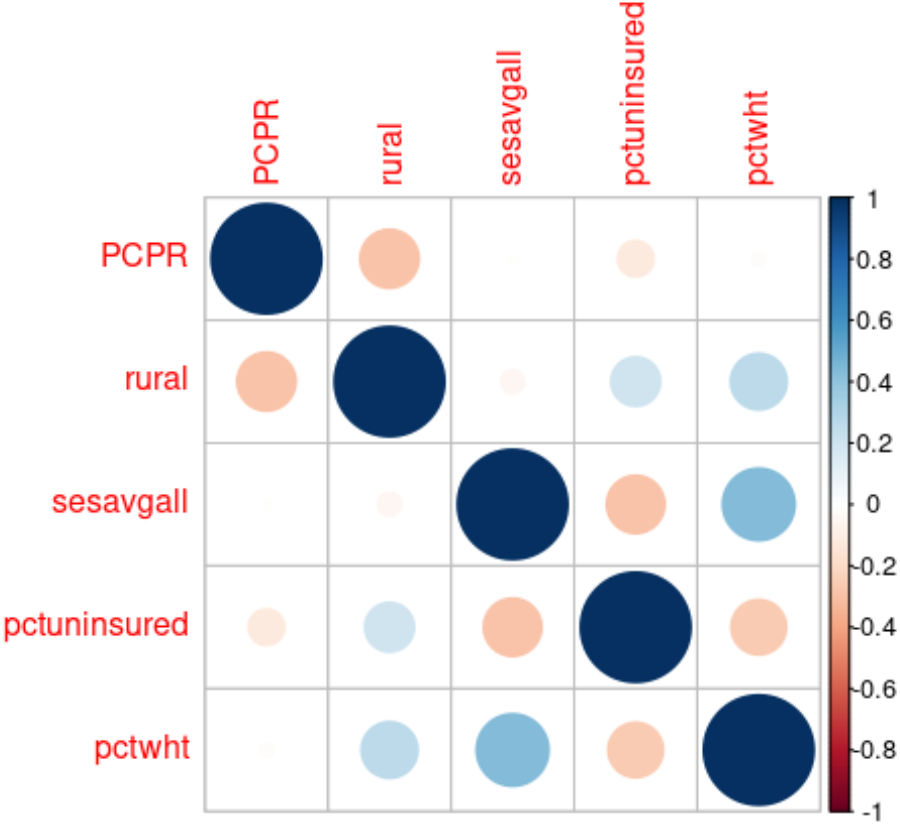
Correlations Between Physician-to-Child Ratio (PCPR), Rate of Uninsured Children, and District Covariates.

While the demographics of a district are somewhat associated with the density of these physicians—districts with more White students have slightly more pediatric providers—rurality has an especially pronounced association (Figure S1). Table 1 shows that pediatric providers are overrepresented in non-rural school districts compared to rural school districts by more than a 2 to 1 margin; the mean rural district has 1.65 doctors per 1,000 children compared to 3.44 doctors in non-rural districts. In fact, over 49% of rural districts have zero pediatric providers, while only 8% of non-rural districts have zero pediatric providers. These patterns are more dramatic among pediatricians, who are overrepresented in non-rural districts by more than a 7 to 1 margin; almost 90% of rural districts have no pediatrician within its boundaries, compared to 38% of non-rural districts. This discrepancy between rural and non-rural districts is further captured in Figure 2 which shows histograms of PCPR for each district. Taken together, rural students have less access to pediatric providers, and this is particularly true for rural places with large non-White populations. In contrast, the rate of uninsured children appears to vary more with sociodemographic characteristics than between rural and non-rural districts (Figure S2).

**Table 1.** Descriptive Statistics, Rural and Non-Rural Districts.

| Measure | Rural Districts |  |  |  | Non-Rural Districts |  |  |  |
| --- | --- | --- | --- | --- | --- | --- | --- | --- |
|  | Mean | SD | Min | Max | Mean | SD | Min | max |
| Pediatrician-to-Child Ratio | 0.11 | 0.58 | 0.00 | 20.94 | 0.79 | 1.38 | 0.00 | 25.10 |
| Physician-to-Child Ratio | 1.65 | 2.71 | 0.00 | 30.30 | 3.44 | 3.20 | 0.00 | 33.46 |
| Rate of Uninsured Children | 7.75 | 6.42 | 0.00 | 43.98 | 5.44 | 4.24 | 0.00 | 36.49 |
| 3rd Grade Achievement | -0.02 | 0.32 | -2.44 | 1.66 | 0.06 | 0.36 | -1.56 | 1.15 |
| Socioeconomic Status | 0.29 | 0.71 | -4.12 | 2.63 | 0.37 | 0.97 | -4.40 | 2.91 |
| Percent White | 80.64 | 23.75 | 0.00 | 100.00 | 66.07 | 28.68 | 0.00 | 99.61 |
| N | 6,421 |  |  |  | 5,875 |  |  |  |

**Figure 2.**
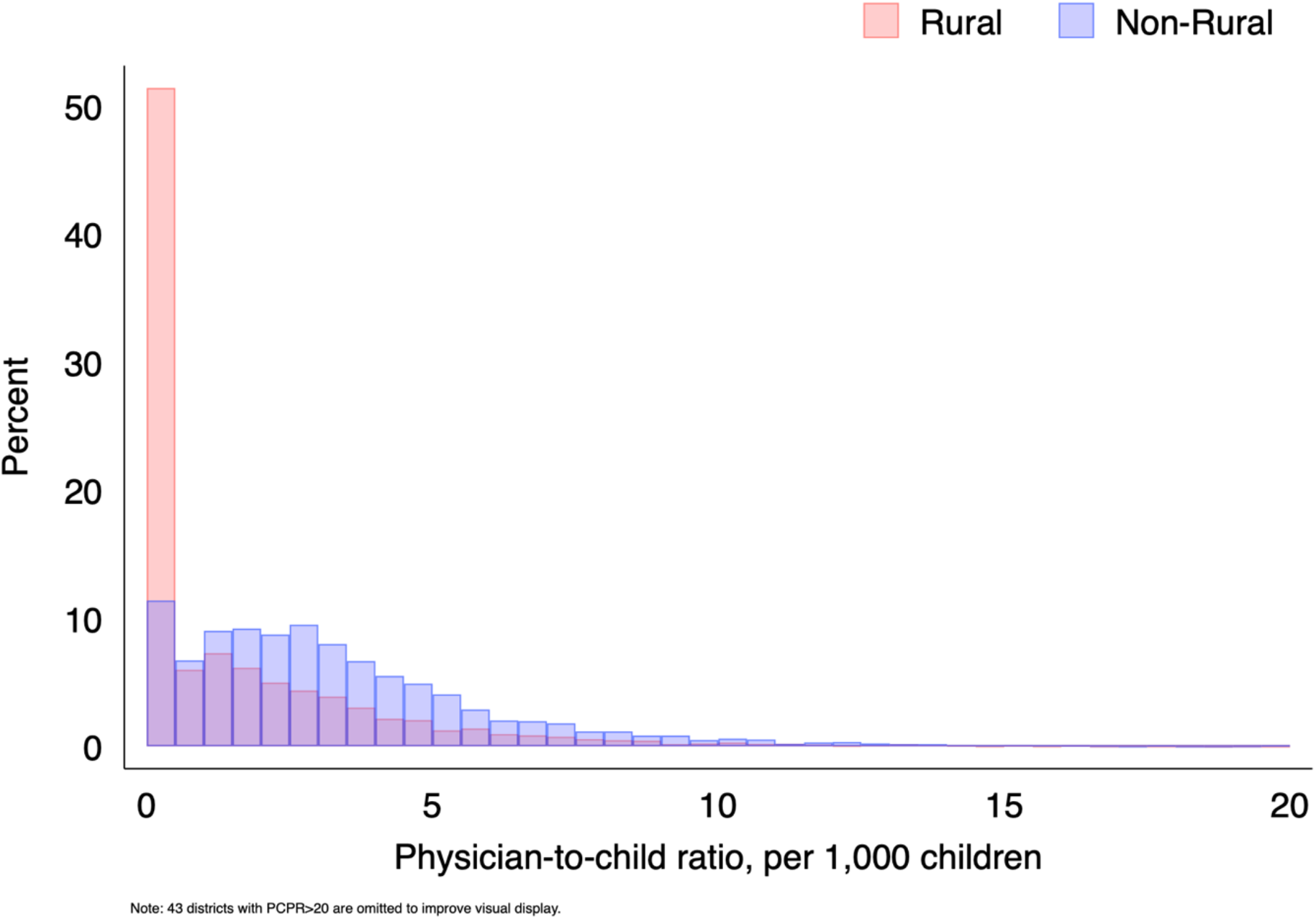
Distribution of Physician-to-Child Ratio (PCPR), Rural and Non-Rural Districts.

We can compare inequality in the distribution of pediatric providers to other kinds of inequality via the Gini coefficient. We calculate a Gini coefficient for PCPR of 0.59. This inequality is even more pronounced when looking only at pediatricians, for whom the Gini coefficient is 0.82. We can compare this to the Gini coefficient for income inequality in the U.S., which was 0.47 in 2018.^37^ Thus, the inequality we observe in the distribution of child-serving doctors is troubling even relative to other pronounced forms of inequality. Collectively, these findings indicate that physician density is (i) a distinctive measure with (ii) a notable level of inequality. We turn now to an analysis of its association with child academic outcomes.

### Associations Between Local Physician Supply and Educational Achievement

Children in districts with more pediatric providers do better in school as measured by achievement tests in third grade. Our baseline result suggests that adding an extra pediatric provider per 1,000 children is associated with a nearly 0.01 SD increase in achievement, while adding an additional pediatrician is associated with a 0.02 increase in achievement (Table 2). Although this is a modest increase, associations are highly heterogeneous; associations are much larger in districts with relatively low levels of PCPR.

**Table 2.** Associations Between Child Health Access and Third Grade Achievement.

|  | Full Data |  | Tertiles |  |  |
| --- | --- | --- | --- | --- | --- |
|  |  |  | (1st) | (2nd) | (3rd) |
| Pediatrician-to-Child Ratio | 0.020 ***<br>(0.002) |  |  |  |  |
| Physician-to-Child Ratio |  | 0.008 ***<br>(0.001) | 0.108 ***<br>(0.029) | 0.021 ***<br>(0.004) | 0.004 ***<br>(0.001) |
| Rate of Uninsured Children | -0.004 ***<br>(0.001) | -0.004 ***<br>(0.001) | -0.002<br>(0.002) | -0.004 ***<br>(0.001) | -0.010 ***<br>(0.001) |
| Socioeconomic Status | 0.216 ***<br>(0.003) | 0.220 ***<br>(0.003) | 0.190 ***<br>(0.007) | 0.223 ***<br>(0.004) | 0.237 ***<br>(0.004) |
| Rural School District | -0.086 ***<br>(0.004) | -0.080 ***<br>(0.004) | -0.077 ***<br>(0.011) | -0.049 ***<br>(0.006) | -0.066 ***<br>(0.007) |
| Percent White | 0.003 ***<br>(0.000) | 0.003 ***<br>(0.000) | 0.003 ***<br>(0.000) | 0.002 ***<br>(0.000) | 0.003 ***<br>(0.000) |
| State fixed effects | Yes | Yes | Yes | Yes | Yes |
| Constant | -0.022 ***<br>(0.003) | -0.039 ***<br>(0.004) | -0.071 ***<br>(0.010) | -0.058 ***<br>(0.009) | -0.006<br>(0.006) |
| N | 12,296 | 12,296 | 4,099 | 4,099 | 4,098 |
| R-squared | 51% | 51% | 35% | 60% | 59% |

Notes: \* p<0.05, \*\* p<0.01, \*\*\* p<0.001. Standard errors listed in parentheses. Rate of uninsured children and percentage of White students are mean centered.

We illustrate this heterogeneity in two ways. First, we consider analysis in districts when they are split into tertiles. The first tertile represents districts with the lowest supply of pediatric providers (0.46 physicians per 1,000 kids, on average) while the third tertile represents districts with the highest supply (5.73 physicians per 1,000 kids, on average). Adding an extra physician in a district with a high physician-to-child ratio is associated with a marginal increase of just 0.004 SDs whereas the association in underserved districts is nearly 0.11 SDs, an increase of 2700%. In other words, an additional pediatric provider per 1,000 kids in underserved districts is associated with an additional third of a grade level of achievement. Second, we allow for nonlinearity of the association between PCPR and achievement via B-splines (Figure 3). Note that the increase is quite rapid as PCPR increases from near zero. In contrast, gains are much more modest when PCPR is larger (see Figure S3 for an additional illustration of this). These findings hold in both rural and non-rural settings.

**Figure 3.**
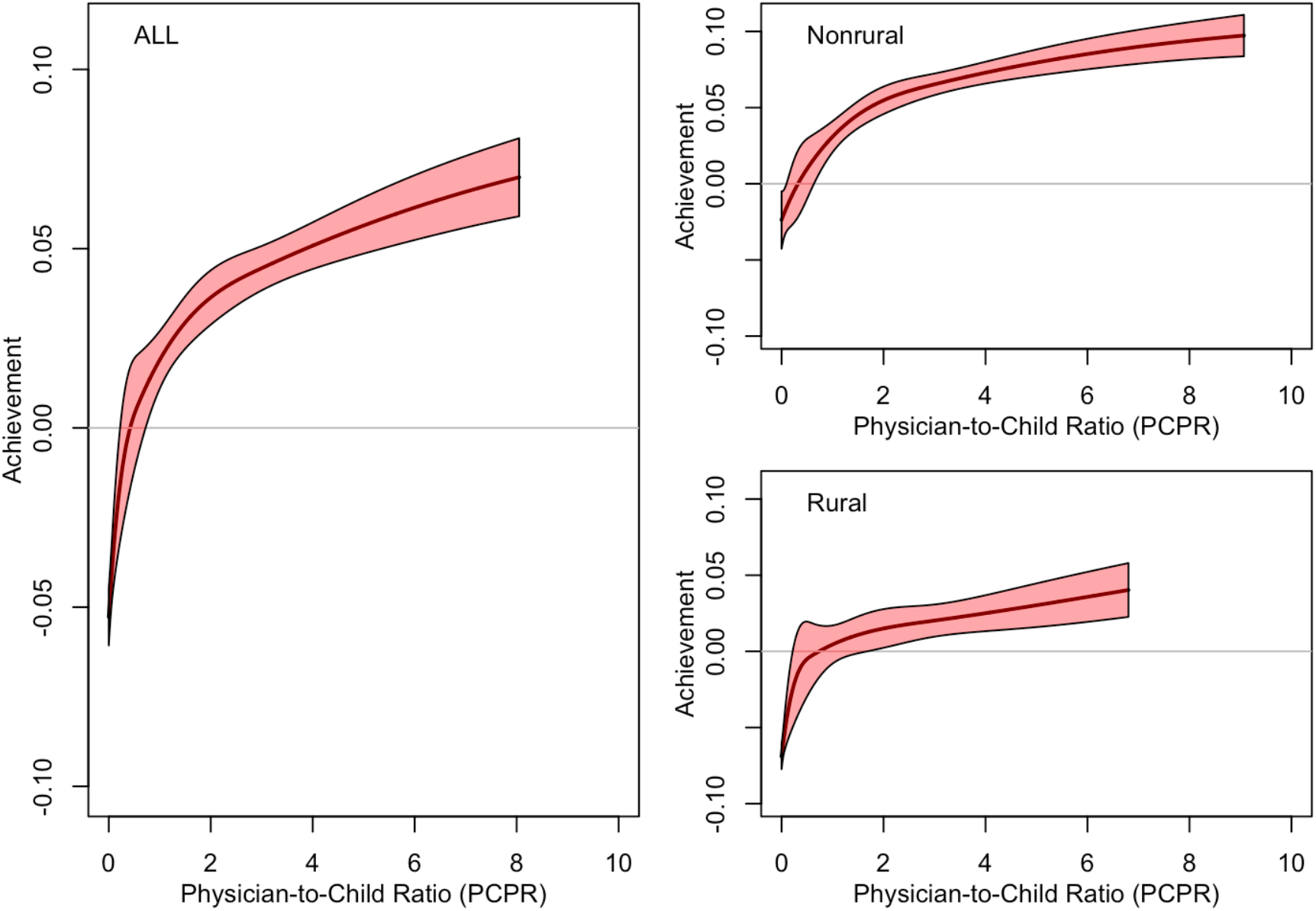
Projected Levels of Achievement as a Function of Physician-to-Child Ratio Using B-Splines.

These findings are robust to a variety of specifications of our model. The overall association is attenuated when we consider all 12,388 districts (column 2, Table S1), but the association among districts in the lowest tertile is nearly unchanged (column 2, Table S2). Because higher physician supply is associated with higher numbers of residency slots in a state, and because these residency slots are often concentrated in urban centers, we remove the 25 largest districts from our model and find that the results are nearly identical to our original model (column 3, Table S1). Results for this specification are completely identical in the lowest tertile because none of the 25 largest districts fall in the lowest third of the PCPR distribution (column 3, Table S2). When we focus solely on controlling for community demographics—dropping state fixed effects and the dummy for rural districts—the association between PCPR and achievement doubles (column 4, Tables S1 and S2). Finally, our simplest model includes just PCPR and the rate of uninsured children, which accounts for 14% of the variance in overall district achievement. The coefficient for the rate of uninsured children becomes much larger in the absence of district SES.

Finally, we examine state-level associations between physician supply and third-grade achievement to discern the extent to which this pattern varies from state to state. Figure 4 depicts the results of these state-level regressions. We observe a positive relationship in every state for which the coefficient is statistically significant; the association in the remaining 17 states cannot be distinguished from zero.

**Figure 4.**
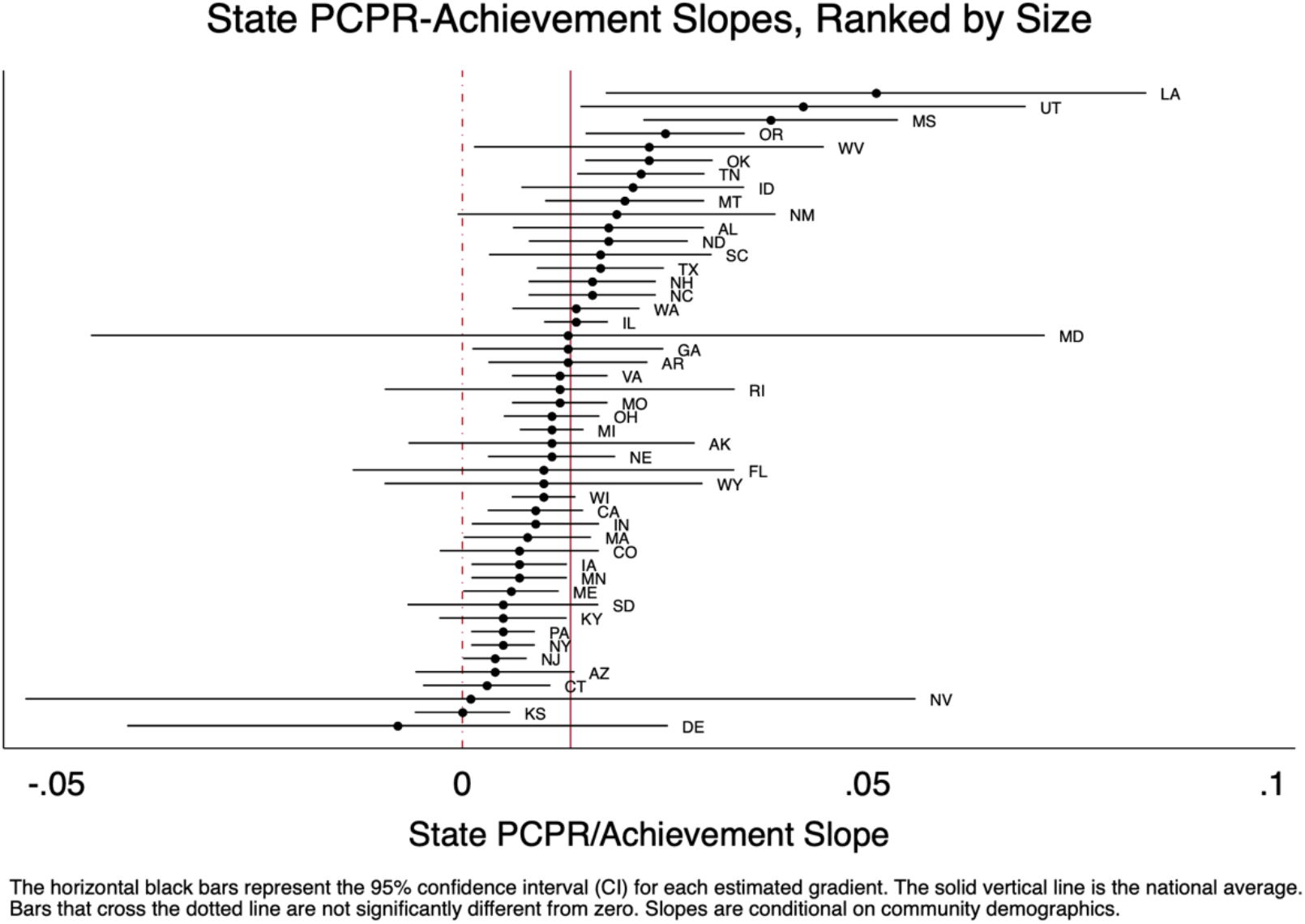
State-Level Associations Between Physician-to-Child Ratio (PCPR) and Achievement.

## Discussion

The distribution of pediatric providers is highly unequal in the United States. In particular, rural students and students of color have lower access to pediatric providers. Shockingly, nearly 90% of rural school districts have no pediatrician within their boundaries and 50% have no pediatric provider at all, highlighting the disparate access to pediatric care experienced by rural students in particular. The density of these physicians in a district is also orthogonal to other features of a district, especially socioeconomic status, and thus merits further attention as a novel feature of the landscape relevant to child development.

Further, we find evidence that the distribution is associated with student achievement. There is a positive relationship between physician-to-child-population ratio and third-grade achievement that operates independently of community socioeconomic status and racial/ethnic composition. Notably, this relationship attenuates in places with higher levels of physician supply, suggesting that it may be possible to generate improvements in early childhood educational opportunity by redistributing the existing physician supply. In other words, we don’t necessarily need to create more pediatric providers—rather, we may simply need to redistribute the ones we already have. Finally, our state-level comparisons demonstrate that this relationship is more salient in some states than others; we hope this finding might spur researchers in states such as Louisiana, Utah, Mississippi, and Oregon to undertake causal analyses using state-level data.

We acknowledge limitations. Some geographic school districts are quite small, and there are a number of feasible scenarios in which families may utilize pediatric care outside of their school district—particularly in small suburban and rural districts that are relatively close to more densely populated areas. Still, health care researchers have wrestled with the fact that there is no obvious geography for health care, especially when seeking to understand the health landscape at the sub-county level.^38^ Because child wellbeing is fundamentally influenced by the health and education landscape in which they live, a district-level measure of physician supply is a potentially important measure for education and health researchers seeking to understand the intersection of these systems. An additional limitation is that the achievement data is drawn from grades 3 through 8, so we cannot speak to the trends in achievement in earlier or later grades, which may differ from what we observe. Moreover, we cannot explain the mechanisms underlying the relationship we see between PCPR and achievement; we are merely describing associations

## Conclusion

While this analysis is not causal, these findings can inform policy in several ways. First, physician training is publicly funded. If communities are reaping uneven and disproportionate benefits from taxpayers’ contributions to America’s medical workforce, policymakers should develop, improve, and monitor policies aimed at distributing pediatric providers in a more equitable way. Second, there have been a number of policy efforts to increase the number of doctors in the U.S. It is important to know if these efforts would perhaps be more effective if they focused on incentivizing physicians to practice in underserved geographic areas. Further investigation of this concave relationship could provide evidence to support the redistribution of the pediatric provider workforce. Finally, expanding medical student loan forgiveness may be an effective way to achieve this redistribution, since research has demonstrated that physicians with more education debt are less likely to serve in health professional shortage areas.^39^

## Data Availability

The test score data and physician location data are publicly available.

https://edopportunity.org/

https://www.cms.gov/Regulations-and-Guidance/Administrative-Simplification/NationalProvIdentStand/DataDissemination

## Supplemental Information

**Table S1.**
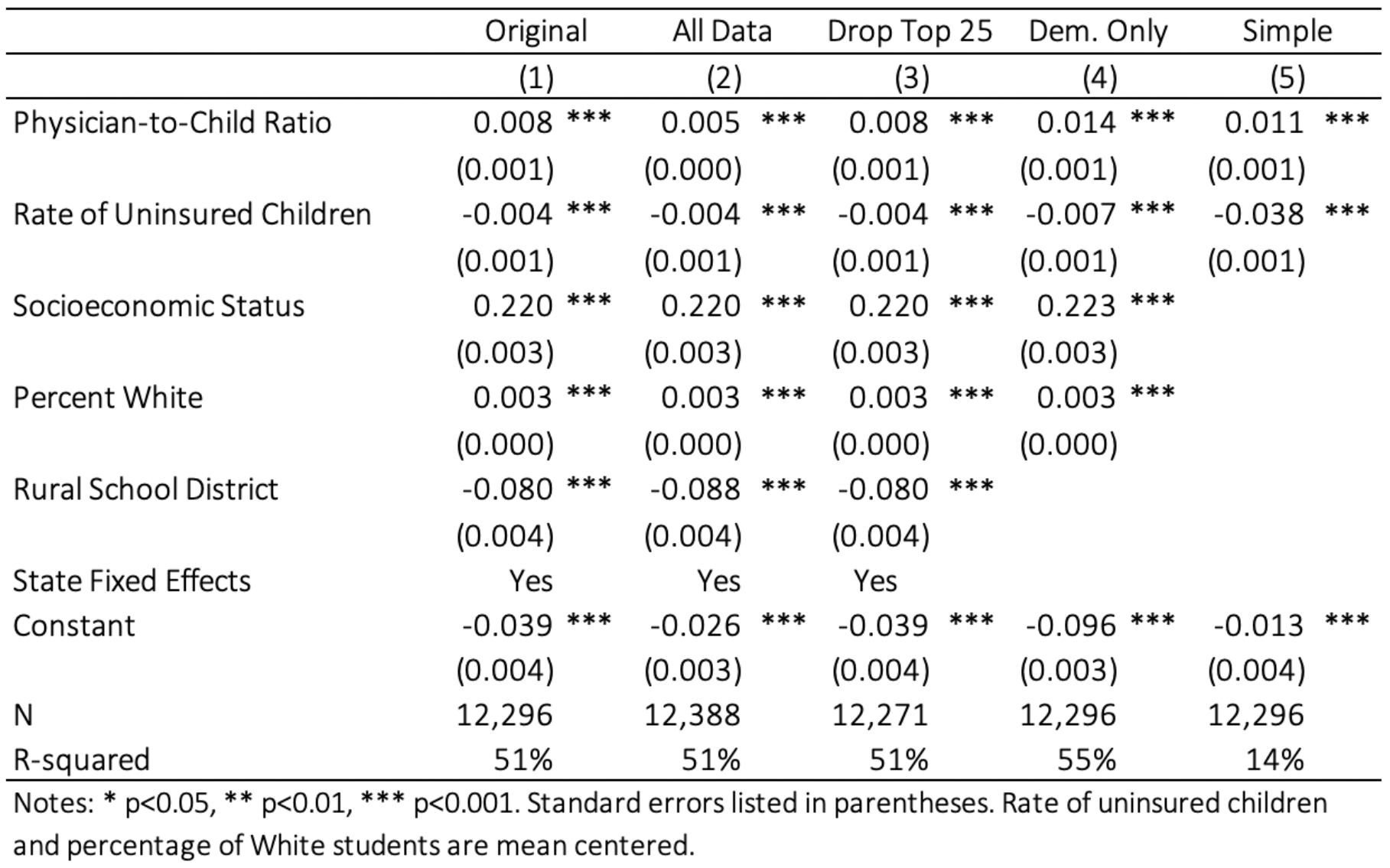
Sensitivity Analyses, All Tertiles.

**Table S2.** Sensitivity Analyses, Lowest Tertile.

|  | Original | All Data | Drop Top 25 | Dem. Only | Simple |
| --- | --- | --- | --- | --- | --- |
|  | (1) | (2) | (3) | (4) | (5) |
| Physician-to-Child Ratio | 0.108 ***<br>(0.029) | 0.106 ***<br>(0.029) | 0.108 ***<br>(0.029) | 0.241 ***<br>(0.030) | 0.065<br>(0.038) |
| Rate of Uninsured Children | -0.002<br>(0.002) | -0.002<br>(0.001) | -0.002<br>(0.002) | -0.004 *<br>(0.001) | -0.037 ***<br>(0.002) |
| Socioeconomic Status | 0.190 ***<br>(0.007) | 0.188 ***<br>(0.007) | 0.190 ***<br>(0.007) | 0.205 ***<br>(0.007) |  |
| Percent White | 0.003 ***<br>(0.000) | 0.003 ***<br>(0.000) | 0.003 ***<br>(0.000) | 0.005 ***<br>(0.000) |  |
| Rural School District | -0.077 ***<br>(0.011) | -0.077 ***<br>(0.011) | -0.077 ***<br>(0.011) |  |  |
| State fixed effects | Yes | Yes | Yes |  |  |
| Constant | -0.071 ***<br>(0.010) | -0.070 ***<br>(0.010) | -0.071 ***<br>(0.010) | -0.149 ***<br>(0.005) | -0.041 ***<br>(0.006) |
| N | 4,099 | 4,130 | 4,099 | 4,099 | 4,099 |
| R-squared | 35% | 35% | 35% | 47% | 11% |

Notes: \* p<0.05, \*\* p<0.01, \*\*\* p<0.001. Standard errors listed in parentheses. Rate of uninsured children and percentage of White students are mean centered.

Figures S1-S3 describe various features of the data. Figure S1 shows PCPR as a function of district SES or racial composition. Figure S2 shows the percentage of uninsured children as a function of district SES or racial composition. In both Figures S1 and S2, attention is paid to the differences between rural and non-rural districts. Figure S3 plots the relationship between -third-grade average achievement and PCPR, conditional on community demographics.

**Figure S1.**
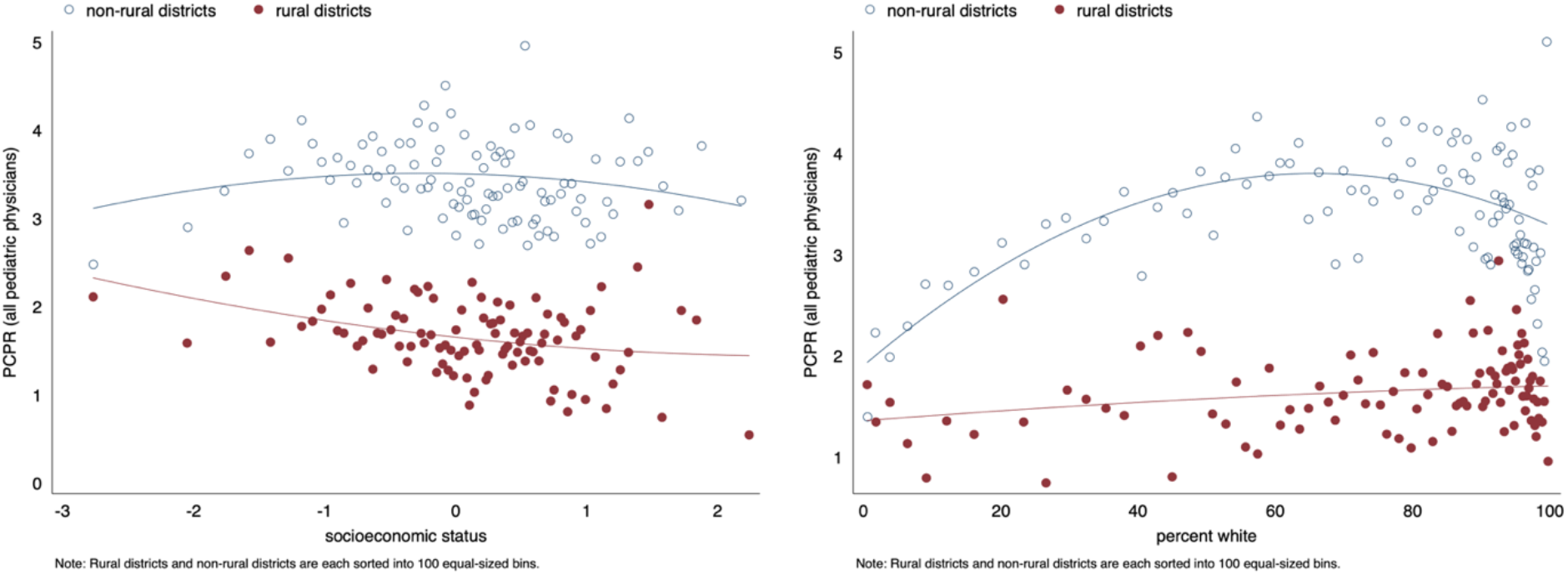
The Relationship Between Physician-to-Child Ratio and Socioeconomic Status (left) & Between Physician-to-Child Ratio and the Percentage of White Students (right).

**Figure S2.**
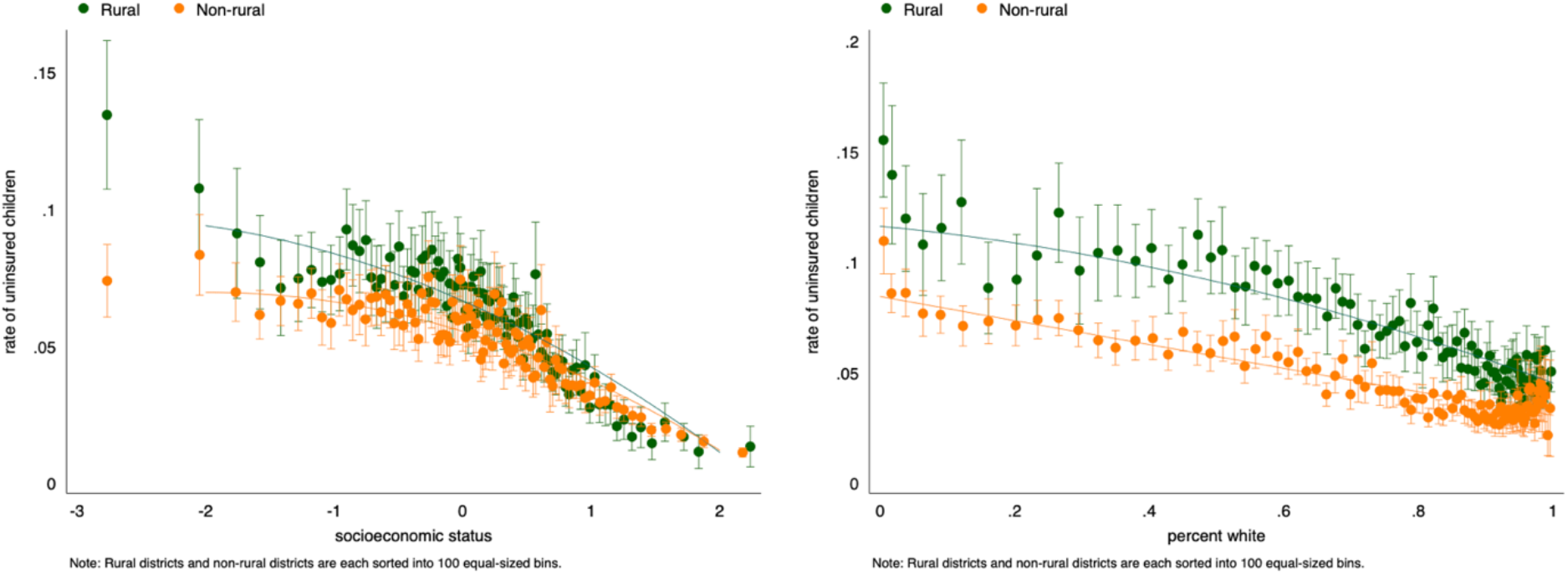
The Relationship Between the Rate of Uninsured Children and Socioeconomic Status (left) & Between the Rate of Uninsured Children and Percentage of White Students (right).

**Figure S3.**
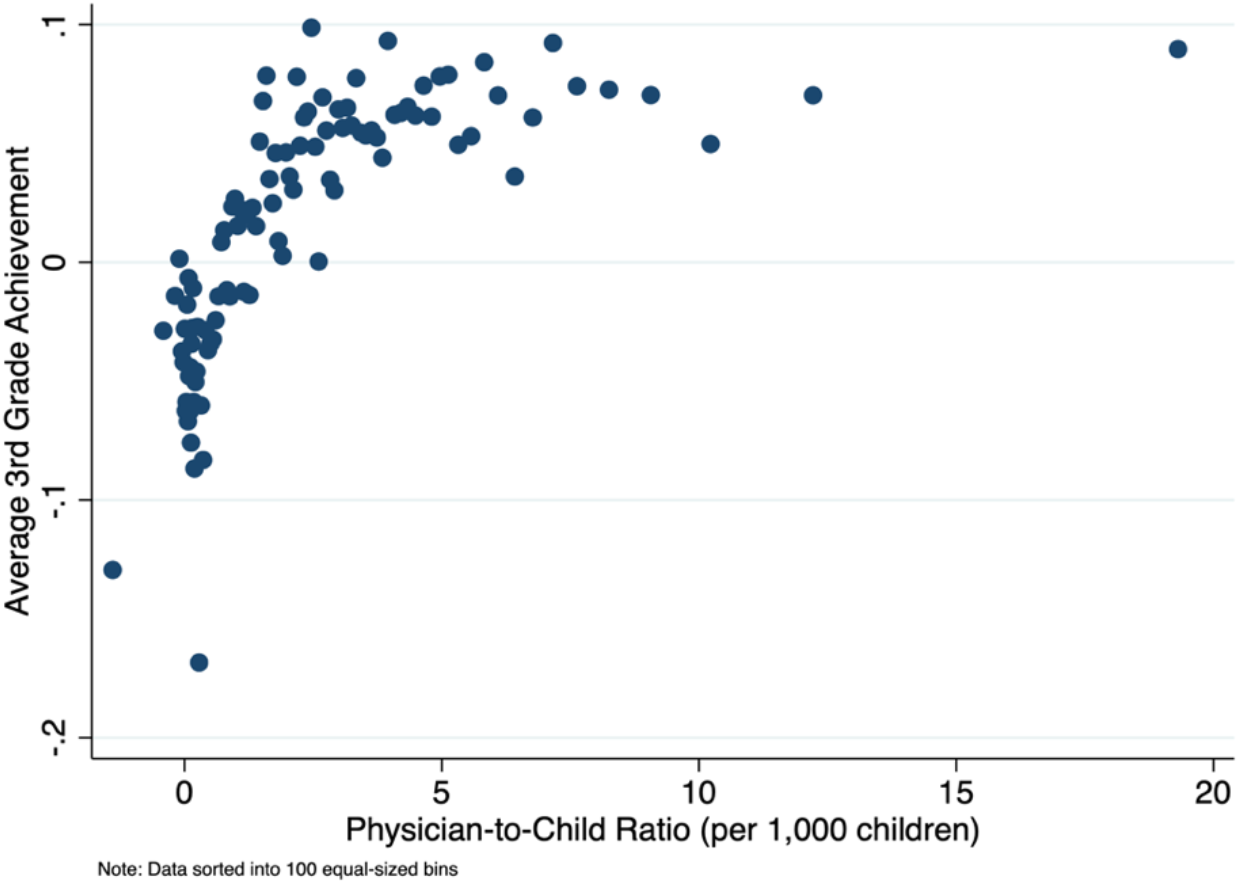
Relationship Between Third Grade Average Achievement and Physician-to-Child Ratio, Conditional on Community Demographics.

## Notes

### Competing Interest Statement

The authors have declared no competing interest.

### Funding Statement

Jessica Boyle is supported by the Institute of Education Sciences, U.S. Department of Education, through Grant R305B140009 to the Board of Trustees of the Leland Stanford Junior University, and by the Robert Wood Johnson Foundation through the Health Policy Research Scholars fellowship award.

### Author Declarations

This was not human subject research. This secondary data analysis is aggregated at the school-district level.

